# Employment Trajectories During the Menopause Transition: Experiences of Women with Early and Surgical Menopause

**DOI:** 10.1101/2024.09.17.24313818

**Authors:** Darina Peycheva, Bozena Wielgoszewska, Paola Zaninotto, Andrew Steptoe, Rebecca Hardy

**Author notes:** Corresponding author: UCL Institute of Epidemiology and Health Care, 1-19 Torrington Pl, London, WC1E 7HB, UK. E-mail address (D. Peycheva).

## Abstract

This study examines the labour market trajectories of women experiencing early and surgical menopause over a 10-year period surrounding their final menstruation or surgery, representing for most women the menopause transition. We also investigate the potential mediating role of hormone therapy (HT) in early postmenopause (within 5 years of menopause onset or surgery) in these relationships. Using data from the English Longitudinal Study of Aging (ELSA), we employ sequence and cluster analysis, followed by mediation analysis. Our findings indicate that women with early menopause, compared to those who undergo menopause at 45 or older, are less likely to have flexible working arrangements (part-time work or self-employment) compared to full-time work during this sensitive period. However, they are equally likely to exit the labour market as to work full-time, indicating distinct experiences. Surgical menopause, compared to natural menopause, is associated with an increased risk of labour market exit, particularly for women aged 45 or older at the time of surgery, potentially due to higher comorbidities and increased age-related precarity. HT use appears to mitigate the risk of labour market exit for women with both early and surgical menopause, although other factors (e.g., health) may play a more significant role in this transition out of work. We advocate for further research on the impact of early and surgical menopause on women’s labour market circumstances and for workplace policies that consider the diverse experiences of women with early and surgical menopause, including their increased risk of chronic conditions.

## 1. Introduction

The onset age of menopause is widely regarded as a key marker of women’s health (Mishra et al., 2024). Notably, menopause before the age of 45, also known as early menopause, is associated with an increased risk of cardiovascular disease, osteoporosis, and premature mortality (Gold, 2011). Women with early menopause are also at higher risk of experiencing more frequent, severe, and prolonged hot flushes and night sweats, as well as depressive symptoms (Paramsothy et al., 2017). Premature morbidity and symptomatic menopause can be even more pronounced in women who undergo early surgical menopause or premenopausal hysterectomy, where ovarian function ceases abruptly or menopause occurs earlier due to disrupted ovarian blood flow (Wilson et al., 2016; Zhu et al., 2020).

Despite extensive research on the health consequences of early and surgical menopause, there is limited evidence on their impact on other aspects of women’s lives, such as labour market experiences. This is important as women constitute a growing proportion of the workforce, and many experience menopause while in work. Surgical menopause often occurs in younger women (Sarrel et al., 2016), and these experiences could impact their work opportunities and trajectories. Previous studies on menopause and women’s labour market circumstances have primarily focused on symptoms during the menopause transition in midlife women (Brewis et al., 2017; Atkinson et al., 2021), as do workplace policies related to menopause (UNISON, 2019; Department for Work and Pensions, 2022). While these aspects are important, insights into the overlooked role of early and surgical menopause are needed, as they may significantly impact women’s labour market participation beyond symptomatic menopause. Understanding women’s work circumstances during this sensitive period is crucial for supporting them through this transition and protecting both their health and work outcomes.

Using data from the English Longitudinal Study of Aging (ELSA), a nationally representative survey of people aged 50 and above in England (Steptoe et al., 2013), we examine the labour market trajectories of women experiencing early menopause (before 45 years) and surgical menopause (bilateral oophorectomy and hysterectomy with preserved ovaries^1^) during a 10-year period that encompasses their final menstruation or surgery. This period represents the menopause transition for most women (Harlow et al., 2012). Additionally, we explore the potential mediating role of hormone therapy in early postmenopause (within 5 years of menopause onset or surgery) in the relationship between early and surgical menopause and women’s labour market trajectories. HT is often used by women who undergo surgery and for the treatment of menopause symptoms, and it could therefore be beneficial in allowing women to maintain their employment.

## 2. Previous literature

Research has examined the impact of menopause symptoms on various work outcomes in midlife women, including employment status, working hours, work ability, absenteeism, and leaving work. Cross-sectional and longitudinal studies consistently found that menopause symptoms—vasomotor, like hot flushes, night sweats, and resulting sleep disturbance, psychological and somatic symptoms—affect work circumstances (e.g., D’Angelo et al., 2023; Bryson et al., 2022; Evandrou et al., 2022). The bidirectionality of the association between menopause symptoms and the workplace environment has also been acknowledged; menopause symptoms can be exacerbated by work-related and organisational factors (e.g., D’Angelo et al., 2023; Hardy et al., 2018; Jack et al., 2014).

How menopause status (pre-, peri-, post-menopause) affects work outcomes remains less clear, according to the few studies that have explored these relationships. Most studies did not find evidence of an association between menopause status and work outcomes (Jack et al., 2014; Hardy et al., 2018; Evandrou et al., 2022). However, Hickey et al. (2017) reported that postmenopausal female employees had a lower intention to leave the labour force compared to pre- and peri-menopausal women. Mvundura (2007), using panel data from the American National Longitudinal Survey of Young Women, showed that as women transitioned from premenopause to natural menopause, their labour force participation increased. Postmenopausal women also faced more physical limitations than perimenopausal women, and surgical menopause was associated with additional limitations in daily activities compared to natural menopause (Mvundura, 2007). These findings are consistent with other studies that highlight how women may exert extra effort at work to conceal vulnerabilities related to menopause, but also financial insecurity, caring responsibility, and coexisting health issues (Evandrou et al., 2022; Gartoulla et al., 2016; Fenton & Panay, 2014).

To our knowledge only four studies investigated the role of earlier menopause on work (Bryson et al., 2022; Saarien et al., 2024; Conti et al., 202 Vincent et al., 2024).

Bryson et al. (2022), using data from the Britain’s 1958 National Child Development Study showed that early natural menopause was associated with a large (9 percentage point) reduction in employment rates (full-time or part-time) once women reached their fifties. However, early natural menopause had no effect on women’s full-time employment rates, suggesting an association between early menopause and lower rates of part-time work.

Saarien et al. (2024), using data from the 1966 Northern Finland Birth Cohort linked to disability and unemployment records, studied the impact of menopause before 46 years on work ability, work participation, and disability benefits. The study showed earlier menopause was associated with poorer self-perceived work ability, lower work participation, and a higher disability pension rate in subsequent years.

Another recent study of administrative health and economic records in Norway and Sweden found that menopause-related complaints influenced women’s labour market participation and earnings (Conti et al. 2024). In the sample of women reporting these complaints before age 45, there was no statistically significantly effect on the likelihood of working full-time, hours worked, or earnings in the first two years, but there was a noticeable reduction in labour supply three to four years after the initial complaint, resulting in an almost 20% decrease in earnings.

A recent qualitative study in Australia explored the impact of early and premature menopause on work performance (Vincent et al., 2024). The authors showed that beyond symptoms, the effects on work extend to treatments and coexisting health conditions. Key factors—health, workplace, and personal—intersect to shape career trajectories. Supportive workplaces benefited highly educated women, while less advantaged women experienced altered career paths, including profession changes, reduced hours, or leaving paid employment. Iatrogenic early or premature menopause and complex treatments contributed to interrupted or altered careers. Short-term work impact was also evident due to menopause symptoms associated with natural or iatrogenic menopause or ovarian suppression during cancer treatment.

Studies have noted that hormone therapy (HT) can have a beneficial effect on menopause symptoms, thereby minimizing adverse work outcomes (e.g., Evandrou et al., 2022; Sarrel et al., 1990; 1991). Sarrel (1990; 1991) found that women who were working experienced fewer symptoms at menopause and were better able to cope with work on HT treatment compared to the pre-treatment phase when symptoms were first assessed. Evandrou et al., (2022) found that the association between severe symptoms and the likelihood of exiting employment or reducing working hours varied among HT users. Specifically, a stronger association was observed between severe symptoms and remaining in employment while reducing working hours among HT users. Mvundura (2007) showed that HT use increased the labour force participation of women who had surgical menopause, despite no evidence of a differential impact for women with natural menopause. However, there was no evidence of a differential impact on the number of hours worked or the likelihood of full-time and self-employment in their study. Other studies have shown that midlife women who stopped HT were more likely to leave their jobs than those who continued it (Daysal & Orsini, 2014) or that HT modifies the relationships between type and age of menopause and health (Zhu et al., 2020), which may in turn affect labour force participation (Currie & Madrian, 1999).

## 3. Research gaps and study questions

It is crucial to provide a more comprehensive understanding of the impacts of early and surgical menopause on employment. The few studies that have investigated associations between early menopause and work circumstances have highlighted significant implications for women’s working lives. However, to our knowledge, no other study has explored the typical work trajectories for women experiencing early menopause, surgical menopause, or both, during the sensitive period bracketing the final menstruation. Further, understanding the mechanisms by which menopause affects work circumstances could help better support women in their experiences. No study has yet explored the potential mediating role of postmenopausal HT, which is used as a treatment for menopausal symptoms and is recommended for women undergoing early menopause. Hence, HT may mitigate the impacts of early and surgical menopause on work.

We address the following research questions:

- What are the typical work trajectories of women with early onset of menopause, surgical menopause, and both early and surgical menopause?

We expect that women with early menopause (compared to those who undergo menopause at 45 or older) and surgical menopause (compared to natural menopause) experience distinct work trajectories due to their increased risk for premature morbidity and a more symptomatic menopause transition. These women may undergo more frequent work transitions to better accommodate their personal circumstances. These transitions could include shifts from the standard full-time employment to less conventional arrangements, such as part-time or self-employment, potentially offering more flexibility. Other transitions may lead them out of the labour market, such as unemployment or providing care for home and family, or even early retirement. Additionally, some may choose to remain in more flexible work roles if they are already in part-time or self-employed positions, or transition to inactivity from these more flexible work forms, reducing the time spent in work altogether.

If early menopause and surgical menopause interact synergistically, their combined influence on labour market participation could be more significant than expected based on their individual effects alone.

- Does postmenopausal hormone therapy initiated in the early years following the final menstruation mediate the associations between early and surgical menopause and typical work trajectories?

We anticipate that women who initiate HT around the time of their final menstrual period or surgery may experience more stable work trajectories than those who do not. Specifically, they may have fewer shifts in work status (e.g., transitions to more flexible working or labour market exits), potentially due to a reduction in menopausal-related complaints.

For decades, HT has been used to alleviate menopausal symptoms, though its effects on chronic health conditions are still much debated (British Menopause Society, 2023; The North American Menopause Society Advisory Panel, 2022). Research informing current guidelines highlight beneficial effects for chronic health if initiated in the early years following the menopause (within 10 years). While uptake of HT is therefore influenced by the severity of menopausal and health complaints, other factors, which have evolved over time, such as availability, access, current knowledge, and advice on treatment use, family history, personal preferences, and publicity also play a role.

## 4. Materials and methods

### 4.1 Study population

We use data from the first Life History interview in the English Longitudinal Study of Aging (ELSA) (Steptoe et al., 2013), administered in 2007 (Wahrendorf et al., 2019; Ward et al., 2009). ELSA has followed a representative cohort of men and women aged 50 and older in England since 2002, providing a multidisciplinary resource for research and policy on aging. The data is available via the UK Data Service (UKDS; NatCen Social Research, University College London, Institute for Fiscal Studies, 2023). The Life History interview, a sub-study of ELSA, collected retrospective data on participants’ earlier life experiences, including marital, fertility, residence, employment, and health histories, as well as difficult life events.

The analytical sample includes women who had reached state pension age, 60 for women in these cohorts (Bozio et al., 2010) and had undergone natural or surgical menopause. We excluded women who experienced late menopause (after 55 years) due to their limited employment time before state pension age. To examine employment trajectories around the final menstrual period (FMP), we also exclude women not working at the start of the observation period and those with missing employment history data in this period. This focus allows us to observe employment changes related to menopause. A flow diagram of participants’ inclusion and exclusion criteria is presented in Supplementary Figure S1.

### 4.2 Measures

#### 4.2.1 Outcomes

We used employment history data from the ELSA 2007 Life History interview to create individual labour market participation trajectories for the 10-year period bracketing the final menstruation for each woman (5 years before and 5 years after the FMP). This data included information on economic activity, such as paid jobs lasting 6 months or longer and periods of inactivity of 3 months or more, with associated start and end dates. Employment statuses were categorized as full-time employment, part-time employment, self-employment, unemployment, home/family work, retirement, full-time education, and other forms of inactivity (Wahrendorf et al., 2019).

For this analysis, we combined part-time work and self-employment into a single category, termed flexible working arrangements, due to the small proportion of self-employed women (8.4% to 9.8%) and their similarities in flexibility. We also aggregated all forms of economic inactivity, including unemployment, home/family work, retirement, and other inactive states, due to their small individual sizes. Unemployment ranged from 0.2% to 0.5%, while retirement ranged from 0.1% to 5.6% during the study period. We analysed work status information across three categories: full-time employment, flexible working arrangements, and inactivity.

##### 4.2.1.1 Employment trajectories bracketing the final menstrual period

Given that women often experience bothersome menopause symptoms before their FMP or face treatment burdens due to disease diagnoses, we focus on a 10-year period bracketing the final menstruation: 5 years before and 5 years after the FMP. This timeframe applies to all women, regardless of their age at menopause. This period typically corresponds to late perimenopause and early postmenopause (Harlow et al., 2011). For women who have undergone bilateral oophorectomy or hysterectomy, we use the date of surgery as a reference point.

We used sequence analysis to identify work trajectories, capturing patterns in how women transition through different employment states over time. We employed optimal matching (OM) to quantify sequence similarity or dissimilarity (McVicar & Anyadike-Danes, 2002). Using the computed distance matrix, we performed agglomerative hierarchical clustering to group similar sequences. We also explored alternatives like Partitioning Around Medoids (PAM) and combined PAM and hierarchical clustering, considering various group sizes. Our evaluation criteria included clustering quality measures and theoretically relevant solutions (Studer, 2013).

We found an 8-cluster solution most informative for examining employment trajectories in the 10-year period bracketing the final menstruation: continuous full-time employment, part-time employment or self-employment, transition to full-time employment, part-time employment or self-employment, and transition out of the labour market. Due to limited cases in some clusters, we combined conceptually similar ones into 3 groups for analysis (Table 1): 1) Continuous full-time employment or transition to full-time employment, 2) Continuous part-time employment, self-employment, or transition to part-time or self-employment, and 3) Transitions out of the labour market (unemployment, retirement, looking after home or family, other inactivity) (see Supplementary Figure S2).

**Table 1.** Clusters of employment trajectories bracketing the final menstruation, n=1,386.

| Description | n (%) | Dominant states (%) |
| --- | --- | --- |
| Cluster 1: Continuous full-time employment, or transition to full-time employment<br>Continuous FTE<br>Transition from PTE or SE to FTE | 561 (39.6)<br>503 (35.6)<br>58 (4.0) | FTE (98.9)<br>FTE (57.3); PTE or SE (33.4) |
| Cluster 2: Continuous part-time employment, self-employment, or transition to part-time or self-employment<br>Continuous PTE or SE<br>Transition from FTE to PTE or SE | 600 (44.5)<br>548 (41.1)<br>52 (3.5) | PTE or SE (96.6)<br>FTE (51.1); PTE or SE (46.5) |
| Cluster 3: Transitions out of the labour market (unemployment, retirement, looking after home or family, other inactivity)<br>Transition from PTE or SE to inactivity<br>Transition from PTE or SE to inactivity<br>Transition from FTE to inactivity<br>Transition from FTE to inactivity | 225 (15.9)<br>51 (3.5)<br>41 (3.0)<br>74 (5.1)<br>59 (4.3) | PTE or SE (50.7); Inactivity (45.6)<br>PTE or SE (20.2); Inactivity (77.7)<br>FTE (66.3); Inactivity (33.5)<br>FTE (26.8); Inactivity (64.3) |

Note: Unweighted n (weighted %); FTE=full-time employment, PTE=part-time employment, SE=self-employment

#### 4.2.2 Exposures and mediators

##### 4.2.2.1 Early menopause and surgical menopause; Postmenopausal hormone therapy

In the ELSA 2007 Life History interview, women were asked about the year of their last menstrual period and the reason for amenorrhea if they had not experienced a menstrual period in the 12 months preceding the interview. Additionally, they were asked about their history of hysterectomy and bilateral oophorectomy, along with the years of these surgeries. Information on hormone therapy (HT) use was also collected, including details about the start and end years of treatment if they were not currently on HT at the time of the interview.

Women who were on HT before their last menstrual period and reported natural menopause (no obvious reason for amenorrhea lasting at least 12 months) were excluded from this analysis because it was not possible to assign an accurate date of menopause. Additionally, women with other reasons for the absence of menstruation (such as chemotherapy, contraceptive, or other medications) or whose menopause status could not be determined due to conflicting or insufficient information were also excluded. This study uses data from women with natural or surgical menopause, classified as follows:

♣ Early Menopause: Women who experienced natural or surgical menopause before the age of 45. Women who underwent menopause at 45 years or older were classified as non-early menopausal.

♣ Surgical Menopause: Women who had undergone bilateral oophorectomy (with or without hysterectomy) or hysterectomy (with ovarian conservation) before their last menstrual period. If they had no menstrual periods in the 12 months preceding the interview and no obvious physiological reasons for the amenorrhea, they were classified as naturally menopausal.

♣ Postmenopausal Hormone Therapy in early postmenopause: Women with natural or surgical menopause who started HT within 5 years of their FMP or surgery. This includes women with surgical menopause who were already on HT before their surgery, as it is advised to continue HT after surgery (British Menopause Society, 2021). Women who initiated HT after 5 years of FMP or never used it were classified as having no hormone treatment during the study period^2^.

Given that current guidance recommends initiating HT early in menopause (within 10 years) and that our study period covers 5 years after the FMP, we focus on initiation within the five years since FMP. In this analysis, postmenopausal HT is treated as potential mediator in the relationship between early and surgical menopause and the employment trajectories of women around the time of their FMP.

#### 4.2.3 Covariates

This analysis accounts for a wide range of additional measures of childhood and adulthood experiences preceding the onset of the 10-year timeframe of our study, which are known to influence both the experience of menopause and employment opportunities and trajectories (e.g. Mishra et al., 2019; Wahrendorf et al., 2017).

The childhood characteristics included: adverse childhood experiences (‘no ACEs’ vs ‘any ACEs’), self-rated childhood health (‘excellent or good’ vs ‘fair, poor, or variable’), and childhood smoking (‘no’ vs ‘yes’). Childhood is defined as ‘before age 16’. ACEs included experiences of parental death or separation, parental substance misuse, parental conflict, parental abuse, or parental unemployment lasting longer than 6 months. We also accounted for age at menarche (‘aged 12 or older’ vs ‘aged 11 or younger (early menarche)’.

The adulthood characteristics included: education (‘some education’ vs ‘no educational qualifications’), previous labour market participation (proportion of time spent working from age 15, end of compulsory school age, until the onset of the period of observation), and nulliparity (‘no live born children’ vs ‘live born children’).

Additionally, we included several circumstances at the onset of the period of observation (the start of the 10-year period of our study): children under 18 (‘no’ vs ‘yes’), ill health (‘no’ vs ‘yes’), living with a partner (‘no’ vs ‘yes’), and accommodation type (‘owned’ vs ‘renter or other’).

This analysis also accounted for chronological age (in years) at the 2007 Life History interview and generation (including ‘Greatest’ born before 1928, ‘Silent’ born between 1928 and 1945, and ‘Boomers’ born between 1946 and 1964’).

### 4.3 Statistical analysis

We first present descriptive statistics of the exposures, mediator, and covariates: means and standard deviations, medians and range for continuous variables, percentages for categorical variables (See Table 2).

**Table 2.**
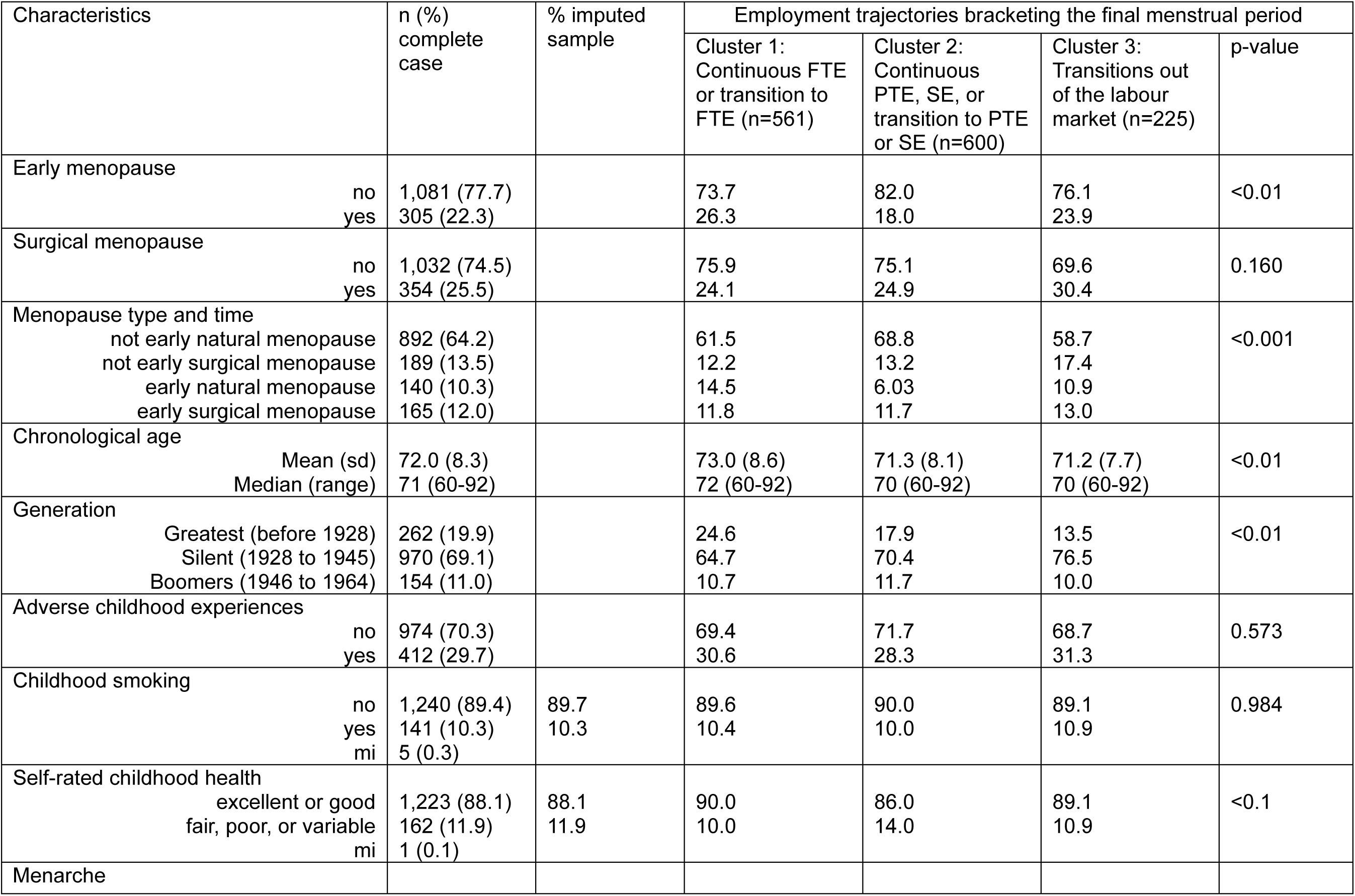
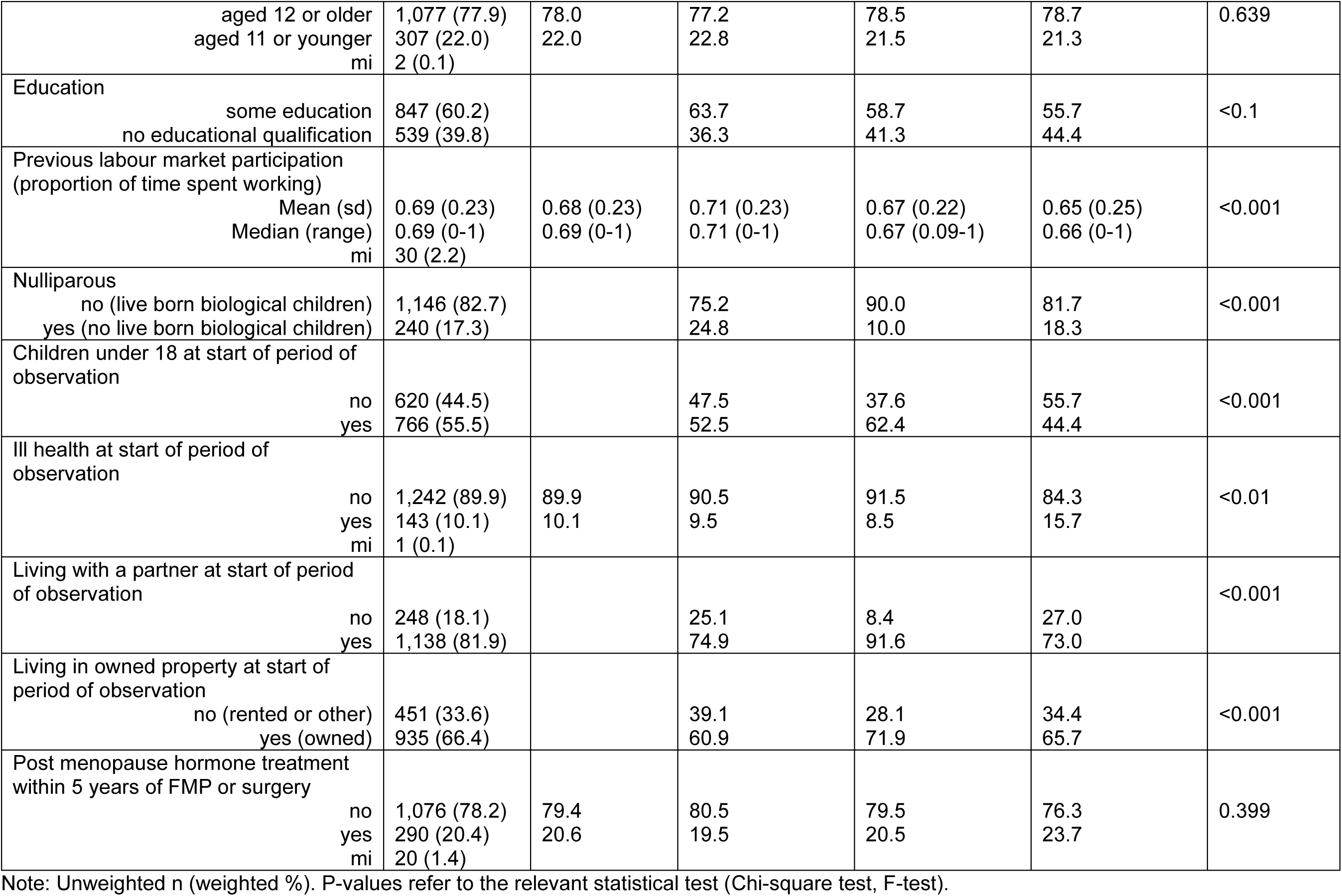
Sample characteristics (mean/percentages) by clusters of employment trajectories bracketing the final menstrual period, n=1,386.

We next investigate how the work trajectories of women, as identified by sequence and cluster analysis (Abbott, 1995; Aisenbrey & Fasang, 2010), are associated with early menopause, surgical menopause, and their combined effects through multinomial regression modelling. The associations between early menopause and surgical menopause with employment trajectory clusters are examined in separate multinomial regression models (early menopause and surgical menopause as independent exposures), followed by a model including an interaction term between early menopause and surgical menopause to examine their combined effect. We use staged covariate adjustment, first accounting for chronological age and generation (model 1), then adding childhood characteristics (model 2), and finally including all covariates, further incorporating adulthood characteristics (model 3). We use survey weight to correct for sampling probabilities and differential nonresponse (Ward et al., 2009). We present relative risk ratios (RRR) and their associated 95% confidence intervals (CI) for all models in Tables 3 and 4.

**Table 3.**
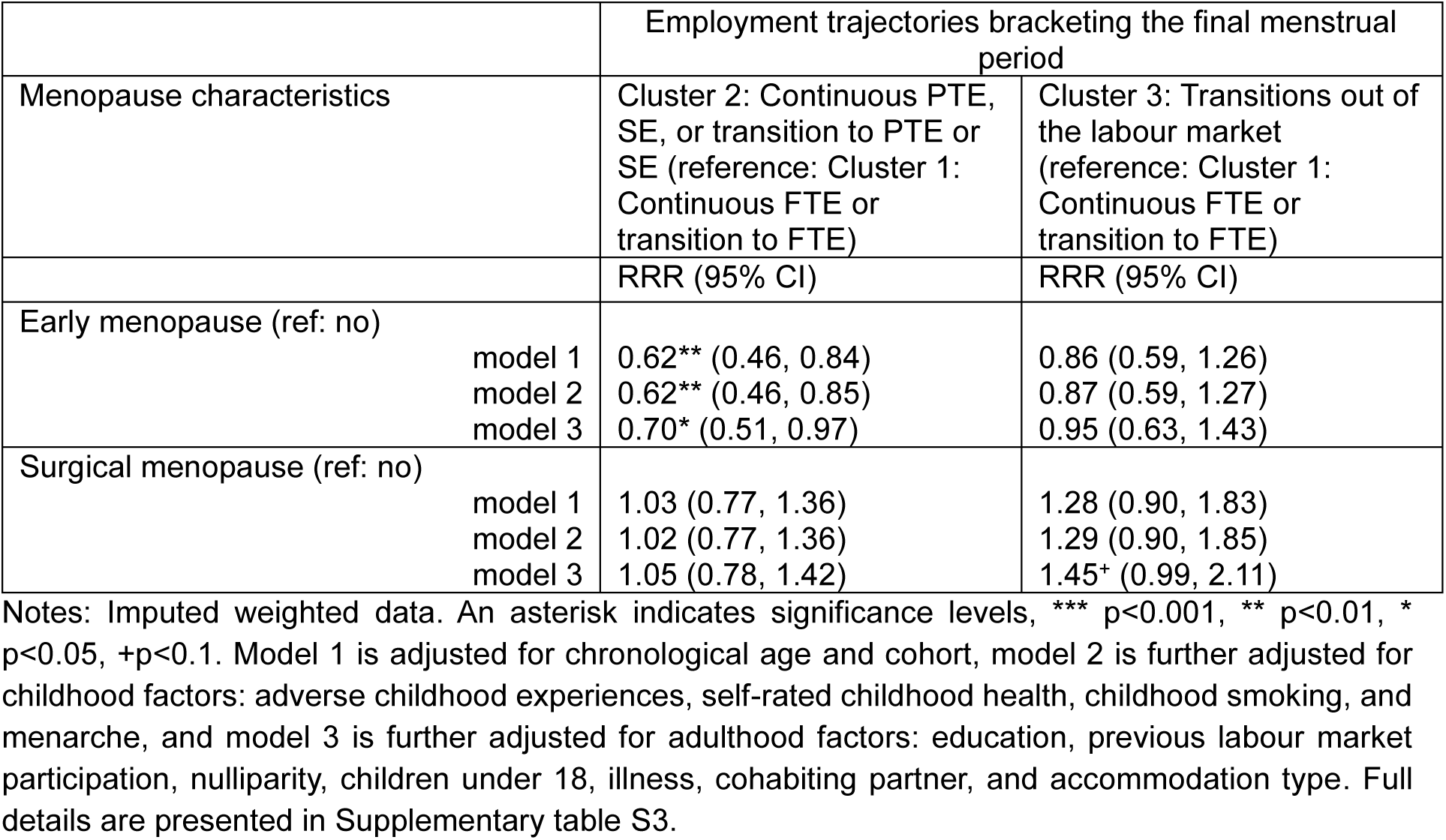
Relative risk ratios (RRRs) and 95% confidence intervals (CIs) of early and surgical menopause and their association with clusters of employment trajectories, n=1,386.

**Table 4.** Relative risk ratios (RRRs) and 95% confidence intervals (CIs) of the combination of early and surgical menopause and their association with clusters of employment trajectories, n=1,386.

|  | Employment trajectories bracketing the final menstrual period |  |
| --- | --- | --- |
| Menopause characteristics | Cluster 2: Continuous PTE, SE, or transition to PTE or SE (reference: Cluster 1: Continuous FTE or transition to FTE) | Cluster 3: Transitions out of the labour market (reference: Cluster 1: Continuous FTE or transition to FTE) |
|  | RRR (95% CI) | RRR (95% CI) |
| Model 1 (Ref. natural menopause at 45 or older) |  |  |
| Surgical menopause at 45 or older | 0.97 (0.67, 1.40) | 1.45 (0.92, 2.27) |
| Early natural menopause | 0.40*** (0.25, 0.62) | 0.82 (0.48, 1.39) |
| Early surgical menopause | 0.87 (0.59, 1.28) | 1.05 (0.64, 1.72) |
| Model 2 (Ref. natural menopause at 45 or older) |  |  |
| Surgical menopause at 45 or older | 0.97 (0.67, 1.40) | 1.45 (0.92, 2.28) |
| Early natural menopause | 0.40*** (0.26, 0.63) | 0.83 (0.49, 1.40) |
| Early surgical menopause | 0.87 (0.58, 1.28) | 1.06 (0.64, 1.75) |
| Model 3 (Ref. natural menopause at 45 or older) |  |  |
| Surgical menopause at 45 or older | 0.95 (0.64, 1.39) | 1.50+ (0.94, 2.38) |
| Early natural menopause | 0.47** (0.29, 0.76) | 0.83 (0.49, 1.41) |
| Early surgical menopause | 0.95 (0.63, 1.43) | 1.28 (0.75, 2.20) |

Notes: Imputed weighted data. An asterisk indicates significance levels, \*\*\* $p < 0.001$ , \*\* $p < 0.01$ , \* $p < 0.05$ , + $p < 0.1$ . Model 1 is adjusted for chronological age and cohort, model 2 is further adjusted for childhood factors: adverse childhood experiences, self-rated childhood health, childhood smoking, and menarche, and model 3 is further adjusted for adulthood factors: education, previous labour market participation, nulliparity, children under 18, illness, cohabiting partner, and accommodation type. Full details are presented in Supplementary table S4.

We quantify the mediating role of postmenopause hormone therapy in the associations between early menopause and surgical menopause with clusters of employment trajectories using causal mediator analysis with inverse odds weighting (IOW) (Tchetgen, 2013; Nguyen et al., 2015), implemented in multinomial regression models. The odds are obtained using a logistic regression model for the exposure given the mediator and confounding variables, accounting for sampling probabilities and differential non-response. The weight is computed by taking the inverse of the predicted odds for each observation in the exposed group; the unexposed group is assigned an IOW of 1. We estimate the total effect (TE) multinomial regression model of the outcome conditional on the exposure and confounding variables, accounting for sampling probabilities and differential non-response. We estimate the natural direct effect (NDE) via a multinomial regression model of the outcome conditional on the exposure and confounding variables, using the IOW multiplied by the survey weight (accounting for sampling probabilities and differential non-response). We calculate the natural indirect effect (NIE) via the mediator by subtracting the NDE from the TE and use 200 bootstrap replications to derive bias-corrected CIs for TE, NDE and NIE.

The sequence and cluster analysis were conducted using R version 4.2.2. Descriptive statistics and regression modelling were conducted using Stata 18.

### 4.4 Missing data

We used multiple imputation (MI) with chained equations to address missing data in the covariates (White et al., 2011). The imputation model includes the outcomes, exposures, mediator, and covariates (von Hippel, 2007). The proportion of missing observations for each variable ranged from 0% to 2.2% (Table 2). We created 20 imputed datasets, and the estimates from each of these datasets were combined to obtain overall estimates using Rubin’s rules (Rubin, 1987).

## 5. Results

Table 2 presents the characteristics of the sample by employment trajectories bracketing the final menstruation. The sample included 1,386 women aged 60 to 92 years (born between 1915 and 1947), with mean age of 72 years at their Life History interview in 2007.

22.3% of women experienced menopause before reaching 45 years: 10.3% experienced early natural menopause and 12.0% had early surgical menopause. In total, 25.5% of women underwent surgical menopause before their final menstrual period^3^.

In terms of childhood experiences, a third of all women (29.7%) had some experience of adversity before they were 16, which included parental separation, family dysfunction, parental abuse, or unemployment. About 10.3% smoked before they were 16 and about 11.9% defined their health in childhood as fair, poor, or variable. 22.0% had early menarche, occurring at age 11 or younger.

About 39.8% of women had no educational qualification. They spent on average about 70% of the time from age 15 (the minimum school leaving age) until the onset of the 10-year period bracketing their final menstruation engaged in work, whether full-time, part-time, or as self-employed. 17.3% were nulliparous (no live born biological children), while more than a half (55.5%) had a child (biological, step, adopted, or foster) under the age of 18 at the start the 10-year period bracketing their final menstruation. 81.9% were living with a partner and 66.4% owned the property in which they resided at the start of the period of observation.

About a fifth of women (20.6%) used hormone therapy within the 5 years following the final menstrual period or surgery^4^.

As shown in Table 1, the first employment trajectory cluster comprised women predominantly engaged in full-time employment or transitioned to full-time employment. 39.6% of women belonged to this cluster. The second cluster included those working part-time, being self-employed, or transitioning to part-time employment or self-employment. 45.5% of women belonged to this cluster. The third cluster consisted of women who transitioned to inactivity, including unemployment, caring for home or family, or other forms of inactivity. This cluster included 15.9% of women in the sample.

Women who worked predominantly full-time or transitioned to full-time work (cluster 1), compared to women in the other clusters, were more likely to experience early natural menopause, have an older mean age of 73 years during the interview, possess some level of education, and were less likely to have biological children (Table 2). Additionally, women belonging to this cluster were more likely to spend more time in work before the onset of the period of observation. They were less likely to live with a partner or own their residence at the onset of the 10-year period.

Women who worked predominantly part-time (cluster 2) were more likely to experience natural menopause after the age of 45 than women in the other 2 clusters. They were more likely to be mothers, to be having dependent children under 18 years old, to live with a cohabiting partner, and own their residence at the onset of the 10-year period surrounding their final menstruation.

Women who exited the labour market during the period of observation (cluster 3) were most likely to undergo surgical menopause. A greater proportion of these women had no educational qualifications and shorter durations of labour market participation prior the period of observation. They were also more likely to report ill health and less likely to have dependent children, under 18, or to cohabit with a partner at the start of the 10-year period surrounding their final menstruation or surgery date.

Table 3 presents the main effects of early menopause and surgical menopause on employment trajectories during the period surrounding the final menstruation. In fully adjusted models (model 3), women with experience of early menopause, compared to those with menopause occurring at age 45 or older, were 30% less likely to work part-time or be self-employed than work full-time (RRR: 0.70, 95% CI: 0.51, 0.97). However, they were as likely to leave the labour market during this period as they were to work full-time (0.95, 0.63, 1.43). Women with experience of surgical menopause, compared to those experiencing natural menopause, were more likely to transition to inactivity and exit the labour market than work full-time (RRR: 1.45, 95% CI: 0.99, 2.11).

Table 4 presents the combined effect of early and surgical menopause on employment trajectories during the period surrounding the final menstruation. While the combination of early and surgical menopause showed an increased, yet not statistically significant, risk of leaving the labour market during the studied period (RRR: 1.28, 95% CI: 0.75, 2.20), an even greater risk was observed for women with surgical menopause at 45 or older (RRR: 1.50, 95% CI: 0.94, 2.38). Women with early natural menopause, compared to those with natural menopause at 45 years or older, were less likely to work part-time than to work full-time (RRR: 0.47, 95% CI: 0.29, 0.76).

We demonstrated associations between early menopause, surgical menopause, and the probability of belonging to a cluster of employment trajectories (Table 3). HT use is hypothesized to be a mechanism linking women’s menopausal experiences to work circumstances during the sensitive period of transitioning into menopause. Surgical menopause was strongly associated with postmenopausal hormone therapy use in this analysis, though no clear evidence of an association between early menopause and hormone therapy use or hormone therapy use and belonging to a specific employment trajectory cluster was observed (Table S5).

Table 5 illustrates the causal effect estimates from the decomposition of the total effect of early menopause and surgical menopause on the likelihood of following certain employment trajectories during the study period. Women with early menopause were 30% less likely to be working continuously part-time, as self-employed, or transition to such employment, compared to women who underwent menopause at 45 or more years (RRR_TE_=0.70, 95% CI_BC_ 0.51, 0.97). No evidence was found for mediation by postmenopausal hormone use (RRR_NIE_= 1.05, 95% CI_BC_ 0.88, 1.26). The difference between the TE and DE estimates was small. independent of hormone therapy use, early menopause was associated with 33% lower likelihood of working part-time compared to working full-time work, (RRR_DE_ 0.67, 95% CI_BC_ 0.47, 0.94). This indicates that factors other than hormone therapy should be considered in understanding the link between early menopause and working full-time rather than part-time.

**Table 5.** Mediation of the association between early and surgical menopause and clusters of employment trajectories by postmenopause hormone therapy, n=1,386.

| Menopause characteristics | Employment trajectories bracketing the final menstrual period |  |
| --- | --- | --- |
|  | Cluster 2: Continuous PTE, SE, or transition to PTE or SE (reference: Cluster 1: Continuous FTE or transition to FTE) | Cluster 3: Transitions out of the labour market (reference: Cluster 1: Continuous FTE or transition to FTE) |
| Effects | RRR (95% CI<sub>BC</sub>) | RRR (95% CI<sub>BC</sub>) |
| Early menopause (ref: no) |  |  |
| Total effect | 0.70 (0.51, 0.97) | 0.95 (0.62, 1.41) |
| Indirect effect (acting through HT) | 1.05 (0.88, 1.26) | 0.79 (0.58, 1.03) |
| Direct effect (unexplained by HT) | 0.67 (0.47, 0.94) | 1.20 (0.73, 1.82) |
| Surgical menopause (ref: no) |  |  |
| Total effect | 1.05 (0.77, 1.46) | 1.45 (1.01, 2.32) |
| Indirect effect (acting through HT) | 0.90 (0.70, 1.17) | 0.73 (0.51, 1.00) |
| Direct effect (unexplained by HT) | 1.17 (0.83, 1.88) | 1.98 (1.28, 3.61) |

Notes: Imputed weighted data. Model is adjusted for chronological age and cohort, childhood factors: adverse childhood experiences, self-rated childhood health, childhood smoking, and menarche, and adulthood factors: education, previous labour market participation, nulliparity, children under 18, illness, cohabiting partner, and accommodation type. Bias-corrected 95% CIs, bootstrapping based on 200 replications.

Despite no evidence of an association between early menopause and transitions out of the labour market (RRR_TE_ 0.95, 95% CI_BC_ 0.62, 1.41), the use of hormone therapy appeared to mediate this relationship (RRR_IE_ 0.79, 95% CI_BC_ 0.58, 1.03) (also known as inconsistent mediation; the IE and DE have different directions, offsetting rather than augmenting each other (Kenny, 2024). This suggests that if all women with early menopause had undergone hormone therapy during the study period, the likelihood of leaving work rather than remaining in full-time employment would have been reduced. Independent of the mediating role of postmenopausal hormone use, early menopause was associated with increased likelihood for leaving the labour market, but the effect was not statistically significant (RRR_DE_ 1.22, 95% CI_BC_ 0.75, 1.86).

Surgical menopause was associated with increased likelihood of exiting the labour market compared to working full-time work (RRR_TE_ 1.45, 95% CI_BC_ 1.01, 2.32). Evidence suggests that postmenopausal hormone therapy use mediated the relationship between surgical menopause and labour market exit. Similar to observations in women with early menopause, if all women with surgical menopause had received hormone treatment, the likelihood of leaving work rather than working full-time would have been lower (RRR_IE_ 0.73, 95% CI_BC_ 0.51, 1.00). Independent of hormone therapy use surgical menopause was associated with a higher likelihood of inactivity compared to full-time employment (RRR_DE_ 1.98, 95% CI_BC_ 1.28, 3.61).

## 6. Discussion

Contrary to our hypothesis, women with early menopause were more likely to work full-time rather than part-time or be self-employed during the menopause transition than women with later menopause. However, they were just as likely to leave the labour market during this period as they were to work full-time.

Our findings—that women with early menopause spent more time in full-time employment compared to more flexible forms of employment—may imply that these women exert extra effort to maintain work stability. Previous studies have shown that postmenopausal women, compared to pre- and peri-menopausal women, are more likely to be working (Myundura, 2007) and have a lower intention to leave the labour market (Hickey et al., 2017). Research has also highlighted menopausal women’s resilience and determination to conceal vulnerabilities related to menopause (Evandrou et al., 2022; Gartoulla et al., 2016; Fenton & Panay, 2014; van den Berg et al., 2010). However, our evidence shows that this maybe particularly the case among women with early menopause.

Their effort could be influenced by the stigma associated with undergoing such an experience at an earlier age than usual, alongside personal circumstances reported previously. It is also possible that the transition into menopause does not significantly disrupt labour market participation, at least in the short term, for some early menopausal women, depending on factors such as the reason for menopause, as well as sociodemographic and work characteristics. Vincent et al. (2024) noted that supportive workplaces were associated with intact career trajectories among educated women with early menopause in their study, which was not the case for less advantaged women with similar menopausal experiences.

However, our results also indicate that women with early menopause were equally as likely as women with later menopause to undergo transitions out of the labour market compared with working in full-time positions, suggesting timing of natural menopause does not impact exits from employment.

These findings align only partly with other studies investigating the impact of early menopause on employment. Bryson et al. (2022) demonstrated in a cohort born in 1958 that early menopause was linked to reduced overall employment rates among women in their fifties, but it had no significant effect on full-time employment rates. In Conti et al.’s (2024) Scandinavian study, menopause complaints before the age of 45 did not significantly impact the likelihood of working full-time or the number of hours worked in the short term. However, three to four years after the initial menopause-related complaint, there was a noticeable reduction in labour supply among this group of women. These differences in the effects on full-time employment could be attributed to generational and population differences between the studies.; Alternatively they might be due to different analytical approaches, e.g., variations in the observation and follow-up periods, differences in how employment outcomes are defined and measured, covariate adjustments. For example, our analysis only considered employment up to 5 years postmenopause, and it may be that the negative impact of early menopause on employment occurs later in the life course.

As expected, women with surgical menopause had a higher risk of leaving the labour market than working full-time during the period surrounding the surgery. The combined effect of early and surgical menopause increased the risk of labour market exit, but this risk was greater for women with surgical menopause at 45 or older. Women with early natural menopause were less likely to work part-time and more likely to work full-time compared to those with natural menopause at 45 or older.

These findings indicate that surgical menopause is a risk factor for labour market exit. This is particularly concerning, not only because of the precariousness that gynaecological surgeries pose for women’s employment, but also because so many women undergo such surgeries. We acknowledge that awareness about surgical menopause and gynaecological surgeries has increased over the years. There is now more support available for women undergoing these procedures than previously (e.g., Surgical Menopause, 2024). However, efforts to understand and normalize the experience of these surgeries and mitigate their impact on women’s work circumstances should be revised and continued.

To our knowledge, this study is the first to quantify the independent effect of surgical menopause on labour market trajectories during the period surrounding the surgery. In their qualitative study, Vincent et al. (2024) noted that beyond symptoms, the effects of early or premature surgical menopause and complex treatments contributed to interrupted or altered careers. Other research focused on the health consequences of surgical menopause (Zhu et al., 2020), which, in line with Vincent’s research, might be important to consider as a pathway to employment disruption among these women.

It is thus not surprising that women 45 or older with experience of surgical menopause may be even more affected compared to those who have the surgery at a younger age. This could be linked to higher comorbidities and increased precarity related to age (e.g., longer recovery times, difficulties in remaining or returning to work, etc.). This aspect warrants further research.

We found no evidence that hormone therapy within the first 5 years of menopause or surgery influenced the likelihood of more flexible work arrangements, such as working part-time or being self-employed, rather than full-time, for women with early or surgical menopause. However, it might have reduced the likelihood of labour market exit. We also observed an increase in the direct effects of early and surgical menopause on labour market exit, independent of hormone use, suggesting that other pathways may play more important role as mediators in this transition.

The finding that hormone therapy in early postmenopause may improve women’s work circumstances by potentially enabling them to work full-time instead of leaving the labour force generally aligns with previous research suggesting that HT may have a beneficial effect on menopausal experiences and could therefore minimize adverse work outcomes (e.g., Evandrou et al., 2022; Daysal and Orsini, 2014). Rather than study the variation in the magnitude and/or direction of the effect of menopausal experiences on employment among women who used HT and those who did not (i.e., HT as an effect modifier) we considered HT as a mediator, i.e., as a potential pathway through which menopausal experiences affect employment. Evidence for mediation can help to identify targeted interventions to support women in the workplace.

Findings such as a potentially reduced risk of labour market exit are important but warrant further research on the role of HT in these relationships. The complexity of HT measurement and analysis makes its effects difficult to compare across studies. Factors such as differences over time in availability, formulations, and advice to use can impact the use of the treatment and affect research. This study also highlights that there might be more important factors that could explain the relationships between menopause type and timing and labour market circumstances. Several studies have indicated that health might be an important factor that plays a role here, either jointly or independently of HT. This potential mediator is worth further research to better understand women’s work circumstances.

### Strengths and limitations

The ELSA Life History interview provides invaluable insights into the complete work histories and comprehensive data on menopause experiences of women in England born in the first half of the 20th century. It allowed us to explore the period surrounding the final menstruation or surgery for each woman, despite differences in the timing of these events. Furthermore, it enabled us to provide robust covariate adjustment across domains such as childhood adversity, health, accommodation, relationships, and fertility histories. Importantly, missing data is minimal and has been addressed through multiple imputation.

Several limitations need to be acknowledged. The information we used was collected retrospectively, which introduces the possibility of recall errors, especially as age increases. However, the reliability and validity of such data improve when event history calendars are used (Belli, 1998), as was the case with the ELSA Life History interview. Changes in job roles, such as moving from a full-time job with more responsibilities to a less demanding full-time job, could have occurred but would have been overlooked in this analysis. Due to sample size limitations, we combined economic activity statuses, such as part-time work and self-employment, into one group. However, these forms of work are conceptually similar in terms of less standard work arrangements, potentially offering more flexibility to manage menopause experiences. Our study’s observation period included time before the final menstruation or surgery, making it challenging to separate the effects of menopause or surgery from other factors, including work-related ones. This may also compromise the mediation analysis in this study, where the temporality of exposure, mediator, and outcome are essential. As menopause is a process, and gynaecological surgeries often involve pre-treatment burdens necessitating the surgical intervention, we deemed the period around the final menstruation or surgery more important for this study than using the final period as a cutoff. We defined surgical menopause as either bilateral oophorectomy or hysterectomy with preserved ovaries. The latter does not lead to immediate menopause and women classified as having surgical menopause in our study may have varying experiences. Women who undergo bilateral oophorectomy may face even more distinct labour market circumstances compared to those who retain at least one ovary after a hysterectomy due to abrupt hormonal changes. A larger sample size is necessary to investigate the differences in experiences between women with bilateral oophorectomy and those with ovarian preservation post-hysterectomy. In this study, postmenopause hormone therapy did not account for HT formulation, dose, duration, or method of administration. Additionally, its use may have been influenced by the severity of complaints, availability, accessibility, current knowledge and guidelines, publicity, and other factors. Measurement error in the mediator can contribute to bias in the causal mediation analysis and underestimate the indirect effects. Lastly, the world of work has evolved over the years, and variations in the timing of reproductive events, including menopause, have been reported. These variations may potentially limit the generalization of findings between birth cohorts.

### Implications

Our study highlights that early and surgical menopause impact women’s labour market trajectories during the transition into menopause. Furthermore, hormone therapy within the early years of the final menstruation may help affected women remain employed. This study strengthens the case for further research and advocates for workplace menopause policies that consider the diverse menopausal experiences of women - the timing and type of menopause. These policies should go beyond mere symptom alleviation and include considerations of their increased risk of chronic conditions. Such overlooked aspects may have significant implications for women’s participation in the labour market and contribute to menopause-related work inequalities.

## Data Availability

All data produced in the present study are available upon reasonable request to the authors

https://doi.org/10.5255/UKDA-Series-200011

## Footnotes

1 Hysterectomy with preserved ovaries does not lead to immediate menopause. However, a hysterectomy without oophorectomy can result in earlier menopause due to reduced ovarian blood supply (Hendrix 2005).

2 No exclusions were made in our study based on the timing of HT initiation. In line with historical availability and usage patterns of HT (Cagnacci & Venier, 2019), HT initiation was uncommon before 1960, with only one woman reporting starting it in 1958. 2% reported starting HT prior to 1980.

3 54.8% of women classified with surgical menopause had a hysterectomy while preserving their ovaries.

4 HT usage was uncommon among women born pre-1928 (3.4%) and rose in later generations—22.7% (Silent Generation, 1928-1945) and 39.6% (Baby Boomers, 1946-1964). This rise aligns with the introduction of oestrogen treatment in the early 1940s and the broader availability of HT in the latter half of the 20th century (Cagnacci & Venier, 2019).

