## Supplemental file for "Employment Trajectories During the Menopause Transition: Experiences of Women with Early and Surgical Menopause"

**Supplementary data**

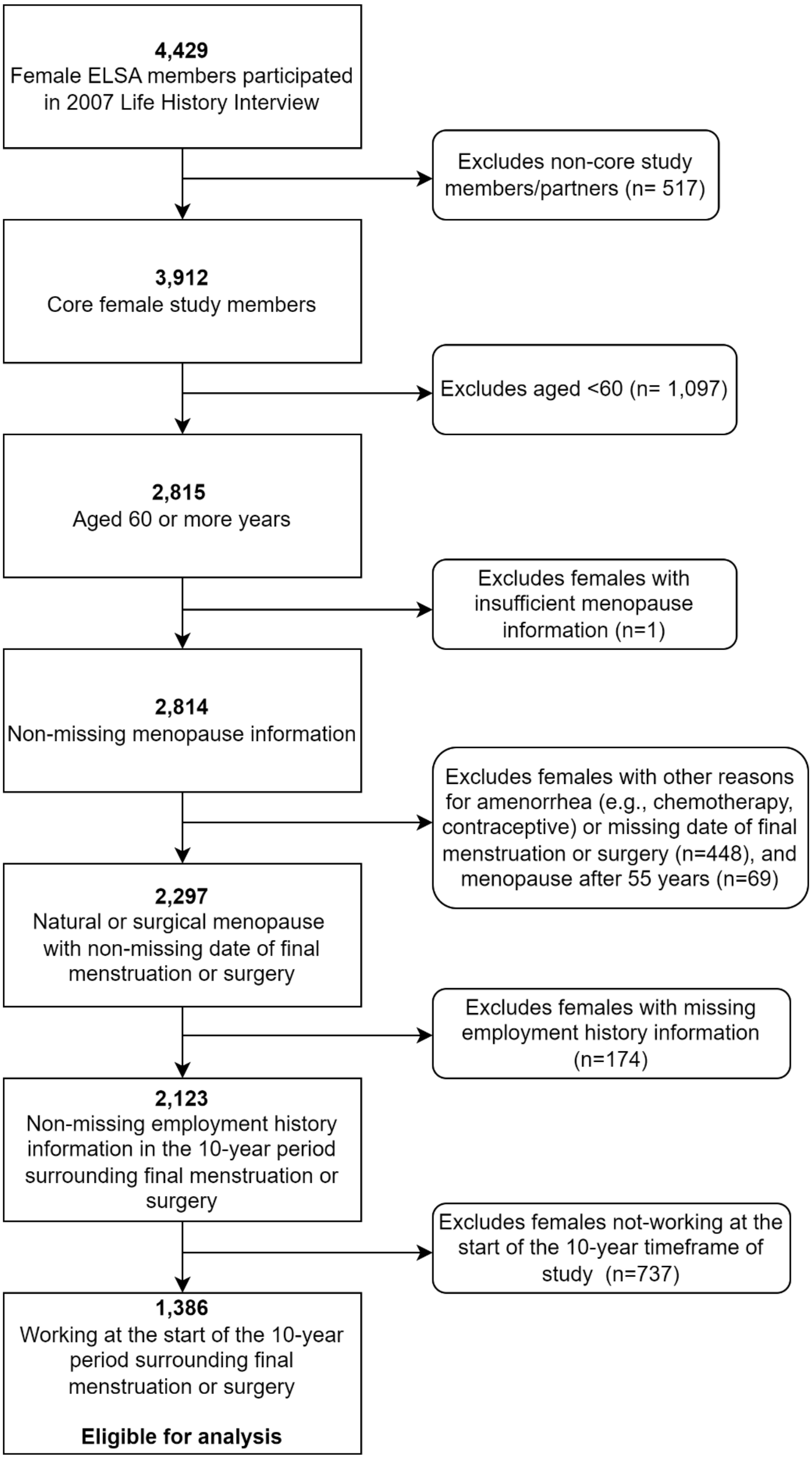

**Figure S1.** Flow diagram of participants' inclusion and exclusion criteria

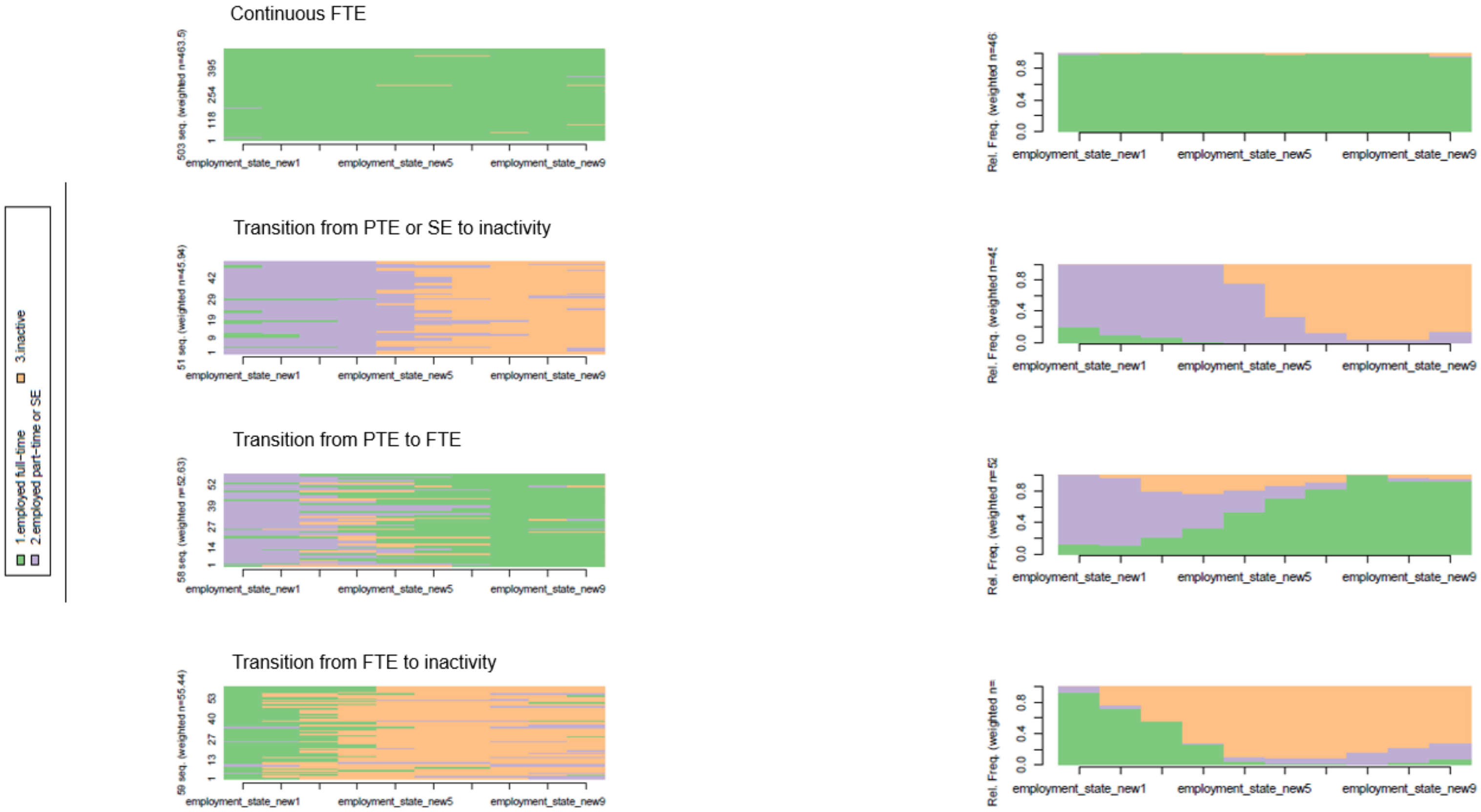

Continuous FTE
Transition from PTE or SE to inactivity
Transition from PTE to FTE
Transition from FTE to inactivity
1.employed full-time
2.employed part-time or SE
3.inactive
employment_state_new1
employment_state_new5
employment_state_new9
Rel. Freq.
0.0
0.4
0.8

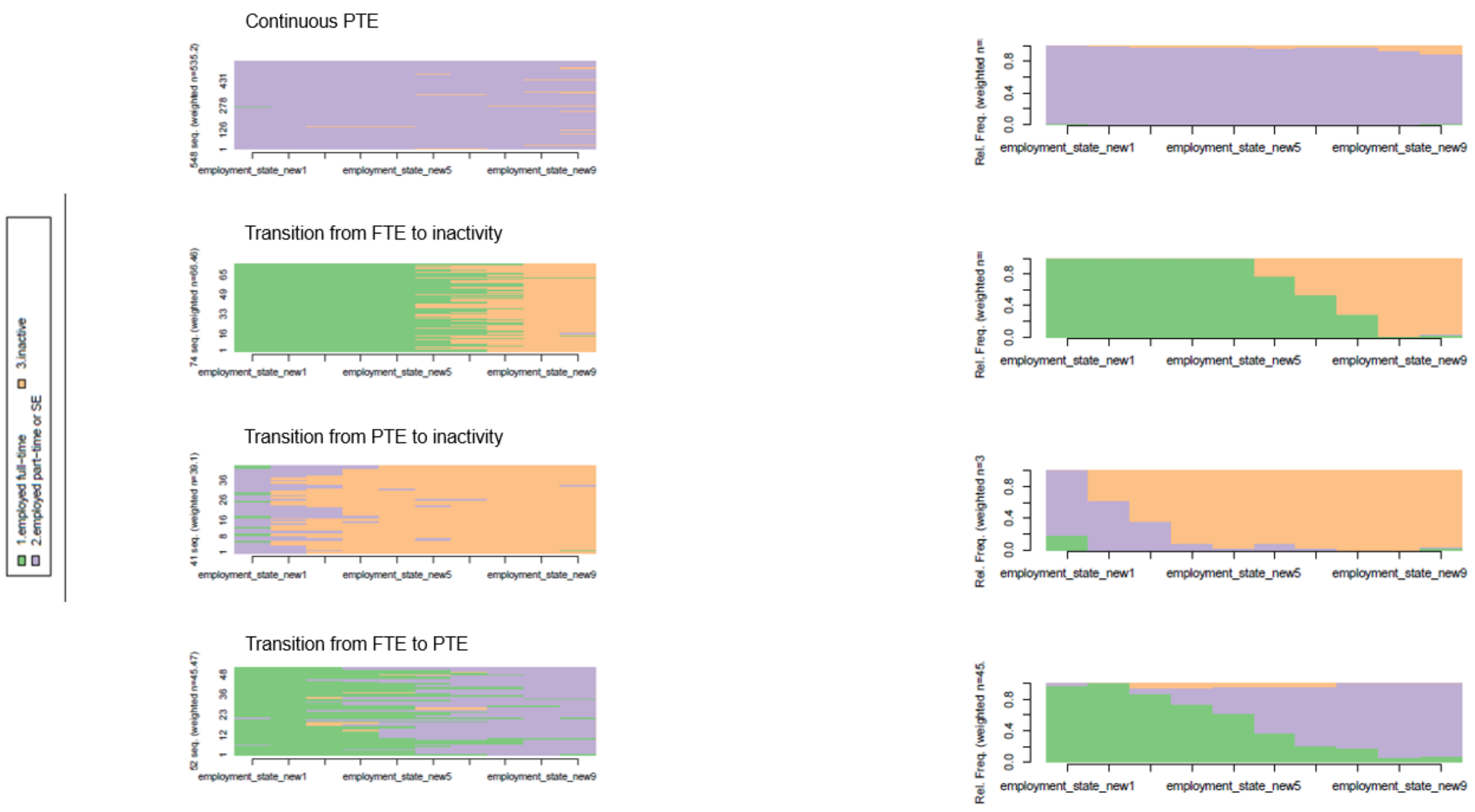

**Figure S2**. Sequence index plots (left) and state distribution plots (right) for the 10-year timeframe of study – 8 cluster solution

**Table S3** Relative risk ratios (RRRs) and 95% confidence intervals (CIs) of early and surgical menopause and their association with clusters of employment trajectories, n=1,386

| | Employment trajectories bracketing the final menstrual period | | | | | |
|---|---|---|---|---|---|---|
| Menopause characteristics | Cluster 2: Continuous PTE, SE, or transition to PTE or SE (reference: Cluster 1: Continuous FTE or transition to FTE) | | | Cluster 3: Transitions out of the labour market (reference: Cluster 1: Continuous FTE or transition to FTE) | | |
| | RRR (95% CI) | | | RRR (95% CI) | | |
| | model 1 | model 2 | model 3 | model 1 | model 2 | model 3 |
| Early menopause (ref: no) | 0.62** (0.46, 0.84) | 0.62** (0.46, 0.85) | 0.70* (0.51, 0.97) | 0.86 (0.59, 1.26) | 0.87 (0.59, 1.27) | 0.95 (0.63, 1.43) |
| Chronological age | 0.97⁺ (0.95, 1.00) | 0.97* (0.95, 1.00) | 0.97* (0.94, 1.00) | 0.99 (0.96, 1.03) | 0.99 (0.96, 1.02) | 0.99 (0.95, 1.02) |
| Generation (ref: Greatest (before 1928) | | | | | | |
| Silent (1928 to 1945) | 1.06 (0.66, 1.70) | 1.03 (0.64, 1.67) | 0.81 (0.49, 1.32) | 1.77⁺ (0.92, 3.43) | 1.77+ (0.92, 3.42) | 1.66 (0.84, 3.25) |
| Boomers (1946 to 1964) | 0.86 (0.40, 1.85) | 0.83 (0.38, 1.80) | 0.61 (0.27, 1.35) | 1.17 (0.41, 3.38) | 1.18 (0.41, 3.38) | 1.17 (0.40, 3.42) |
| Adverse childhood experiences (ref: no) | | 0.86 (0.66, 1.12) | 0.86 (0.65, 1.13) | 0.97 (0.58, 1.63) | 1.02 (0.72, 1.45) | 0.99 (0.70, 1.41) |
| Fair, poor or variable childhood health (ref: excellent or good) | | 1.57* (1.07, 2.30) | 1.64* (1.10, 2.43) | | 1.05 (0.63, 1.76) | 0.98 (0.58, 1.65) |
| Childhood smoking (ref: no) | | 0.92 (0.62, 1.36) | 0.90 (0.59, 1.35) | | 0.97 (0.58, 1.63) | 0.82 (0.48, 1.38) |
| Early menarche (ref: aged 12 or older) | | 0.91 (0.68, 1.23) | 0.93 (0.68, 1.26) | | 0.89 (0.60, 1.32) | 0.89 (0.60, 1.33) |
| Some education (ref: no educational qualification) | | | 0.72* (0.55, 0.95) | | | 0.63* (0.45, 0.89) |
| Previous labour market participation (proportion of time spent working) | | | 0.85 (0.46, 1.57) | | | 0.35** (0.16, 074) |
| Nulliparous (ref: no) | | | 0.50** (0.33, 0.76) | | | 0.65 (0.38, 1.10) |
| Children under 18 at start of period of observation (ref: no) | | | 1.08 (0.81, 1.44) | | | 0.55** (0.38, 0.81) |
| Ill health at start of period of observation (ref: no) | | | 0.88 (0.57, 1.37) | | | 1.66* (1.01, 2.71) |
| Living with a partner at start of period of observation (ref: no) | | | 2.62*** (1.76, 3.89) | | | 0.83 (0.54, 1.29) |
| Living in owned property at start of period of observation (ref: no) | | | 1.27 (0.95, 1.71) | | | 1.19 (0.82, 1.72) |
| Surgical menopause (ref: no) | 1.03 (0.77, 1.36) | 1.02 (0.77, 1.36) | 1.05 (0.78, 1.42) | 1.28 (0.90, 1.83) | 1.29 (0.90, 1.85) | 1.45⁺ (0.99, 2.11) |

| | | | | | | |
|---|---|---|---|---|---|---|
| Chronological age | 0.97* (0.95, 1.00) | 0.97* (0.95, 1.00) | 0.97* (0.94, 1.00) | 0.99 (0.96, 1.03) | 0.99 (0.96, 1.02) | 0.99 (0.96, 1.03) |
| Generation (ref: Greatest (before 1928) | | | | | | |
| Silent (1928 to 1945) | 1.02 (0.63, 1.65) | 1.00 (0.62, 1.62) | 0.78 (0.48, 1.29) | 1.74+ (0.90, 3.38) | 1.75+ (0.90, 3.38) | 1.65 (0.83, 3.24) |
| Boomers (1946 to 1964) | 0.81 (0.38, 1.75) | 0.79 (0.37, 1.72) | 0.58 (0.26, 1.30) | 1.13 (0.39, 3.27) | 1.14 (0.39, 3.29) | 1.15 (0.39, 3.36) |
| Adverse childhood experiences (ref: no) | | 0.85 (0.65, 1.11) | 0.85 (0.65, 1.12) | | 1.00 (0.71, 1.42) | 0.97 (0.68, 1.38) |
| Fair, poor or variable childhood health (ref: excellent or good) | | 1.54* (1.05, 2.56) | 1.63* (1.09, 2.42) | | 1.02 (0.61, 1.71) | 0.96 (0.56, 1.62) |
| Childhood smoking (ref: no) | | 0.86 (0.58, 1.27) | 0.86 (0.57, 1.29) | | 0.93 (0.55, 1.57) | 0.80 (0.47, 1.35) |
| Early menarche (ref: aged 12 or older) | | 0.89 (0.66, 1.19) | 0.91 (0.67, 1.23) | | 0.87 (0.59, 1.29) | 0.88 (0.59, 1.32) |
| Some education (ref: no educational qualification) | | | 0.72* (0.55, 0.95) | | | 0.62** (0.44, 0.88) |
| Previous labour market participation (proportion of time spent working) | | | 0.82 (0.45, 1.50) | | | 0.35** (0.16, 0.74) |
| Nulliparous (ref: no) | | | 0.49** (0.32, 0.74) | | | 0.62+ (0.37, 1.05) |
| Children under 18 at start of period of observation (ref: no) | | | 1.00 (0.75, 1.34) | | | 0.50*** (0.35, 0.74) |
| Ill health at start of period of observation (ref: no) | | | 0.86 (0.55, 1.43) | | | 1.58+ (0.97, 2.58) |
| Living with a partner at start of period of observation (ref: no) | | | 2.72*** (1.83, 4.05) | | | 0.85 (0.55, 1.31) |
| Living in owned property at start of period of observation (ref: no) | | | 1.33+ (0.99, 1.78) | | | 1.22 (0.85, 1.76) |

Notes: Imputed weighted data. An asterisk indicates significance levels, *** $p<0.001$, ** $p<0.01$, * $p<0.05$, +$p<0.1$. Model 1 is adjusted for chronological age and cohort, model 2 is further adjusted for childhood factors: adverse childhood experiences, self-rated childhood health, childhood smoking, and menarche, and model 3 is further adjusted for adulthood factors: education, previous labour market participation, nulliparity, children under 18, illness, cohabiting partner, and accommodation type.

**Table S4** Relative risk ratios (RRRs) and 95% confidence intervals (CIs) of the combination of early and surgical menopause and their association with clusters of employment trajectories, n=1,386

| | Employment trajectories bracketing the final menstrual period | | | | | |
|---|---|---|---|---|---|---|
| Menopause characteristics | Cluster 2: Continuous PTE, SE, or transition to PTE or SE (reference: Cluster 1: Continuous FTE or transition to FTE) | | | Cluster 3: Transitions out of the labour market (reference: Cluster 1: Continuous FTE or transition to FTE) | | |
| | RRR (95% CI) | | | RRR (95% CI) | | |
| | model 1 | model 2 | model 3 | model 1 | model 2 | model 3 |
| (Ref. natural menopause at 45 or older) | | | | | | |
| Surgical menopause at 45 or older | 0.97 (0.67, 1.40) | 0.97 (0.67, 1.40) | 0.95 (0.64, 1.39) | 1.45 (0.92, 2.27) | 1.45 (0.92, 2.28) | 1.50$^{+}$ (0.94, 2.38) |
| Early natural menopause | 0.40*** (0.25, 0.62) | 0.40*** (0.26, 0.63) | 0.47** (0.29, 0.76) | 0.82 (0.48, 1.39) | 0.83 (0.49, 1.40) | 0.83 (0.49, 1.41) |
| Early surgical menopause | 0.87 (0.59, 1.28) | 0.87 (0.58, 1.28) | 0.95 (0.63, 1.43) | 1.05 (0.64, 1.72) | 1.06 (0.64, 1.75) | 1.28 (0.75, 2.20) |
| Chronological age | 0.98$^{+}$ (0.95, 1.00) | 0.97$^{+}$ (0.95, 1.00) | 0.97* (0.94, 1.00) | 0.99 (0.96, 1.03) | 0.99 (0.96, 1.03) | 0.99 (0.96, 1.03) |
| Generation (ref: Greatest (before 1928) | | | | | | |
| Silent (1928 to 1945) | 1.05 (0.65, 1.69) | 1.02 (0.63, 1.66) | 0.79 (0.94, 1.00) | 1.78$^{+}$ (0.92, 3.46) | 1.78$^{+}$ (0.92, 3.45) | 1.68 (0.85, 3.30) |
| Boomers (1946 to 1964) | 0.86 (0.39, 1.86 | 0.83 (0.38, 1.81) | 0.60 (0.27, 1.35) | 1.16 (0.40, 3.35) | 1.17 (0.40, 3.36) | 1.18 (0.40, 3.44) |
| Adverse childhood experiences (ref: no) | | 0.86 (0.66, 1.12) | 0.86 (0.65, 1.13) | | 1.00 (0.71, 1.43) | 0.97 (0.68, 1.38) |
| Fair, poor or variable childhood health (ref: excellent or good) | | 1.53* (1.05, 2.25) | 1.61* (1.08, 2.40) | | 1.02 (0.61, 1.72) | 0.96 (0.56, 1.62) |
| Childhood smoking (ref: no) | | 0.91 (0.61, 1.35) | 0.88 (0.58, 1.34) | | 0.97 (0.57, 1.64) | 0.82 (0.48, 1.38) |
| Early menarche (ref: aged 12 or older) | | 0.92 (0.68, 1.23) | 0.93 (0.68, 1.26) | | 0.88 (0.59, 1.31) | 0.89 (0.60, 1.33) |
| Some education (ref: no educational qualification) | | | 0.72* (0.55, 0.94) | | | 0.62** (0.44, 0.88) |
| Previous labour market participation (proportion of time spent working) | | | 0.85 (0.46, 1.56) | | | 0.35** (0.16, 0.76) |
| Nulliparous (ref: no) | | | 0.49** (0.32, 0.75) | | | 0.63$^{+}$ (0.37, 1.06) |
| Children under 18 at start of period of observation (ref: no) | | | 1.06 (0.79, 1.41) | | | 0.51** (0.35, 0.76) |
| Ill health at start of period of observation (ref: no) | | | 0.87 (0.55, 1.36) | | | 1.58$^{+}$ (0.97, 2.59) |
| Living with a partner at start of period of observation (ref: no) | | | 2.56*** (1.71, 3.82) | | | 0.83 (0.54, 1.28) |
| Living in owned property at start of period of observation (ref: no) | | | 1.27 (0.95, 1.71) | | | 1.19 (0.82, 1.73) |

Notes: Imputed weighted data. An asterisk indicates significance levels, *** p<0.001, ** p<0.01, * p<0.05, +p<0.1. Model 1 is adjusted for chronological age and cohort, model 2 is further adjusted for childhood factors: adverse childhood experiences, self-rated childhood health, childhood smoking, and menarche, and model 3 is further adjusted for adulthood factors: education, previous labour market participation, nulliparity, children under 18, illness, cohabiting partner, and accommodation type.

**Table S5** Associations between early menopause, surgical menopause and postmenopausal hormone therapy (HT); Associations between postmenopausal hormone therapy (HT) and clusters of employment trajectories, n=1,386

| | | Employment trajectories bracketing the final menstrual period | |
|---|---|---|---|
| Menopause characteristics | HT | Cluster 2: Continuous PTE, SE, or transition to PTE or SE (reference: Cluster 1: Continuous FTE or transition to FTE) | Cluster 3: Transitions out of the labour market (reference: Cluster 1: Continuous FTE or transition to FTE) |
| | RRR (95% CI) | RRR (95% CI) | RRR (95% CI) |
| Early menopause (ref: no) | 1.01 (0.71, 1.45)[1] | | |
| Surgical menopause (ref: no) | 4.83*** (3.48, 6.71)[1] | | |
| HT | | 0.93 (0.66, 1.31)[2] | 0.97 (0.61, 1.54)[2] |

[1] Notes: Imputed weighted data. An asterisk indicates significance levels, *** p<0.001, ** p<0.01, * p<0.05, +p<0.1. Model is adjusted for chronological age and cohort, childhood factors: adverse childhood experiences, self-rated childhood health, childhood smoking, and menarche, and adulthood factors: education, previous labour market participation, nulliparity, children under 18, illness, cohabiting partner, and accommodation type.

[2] Notes: Imputed weighted data. An asterisk indicates significance levels, *** p<0.001, ** p<0.01, * p<0.05, +p<0.1. Model is adjusted for menopause type and time, chronological age and cohort, childhood factors: adverse childhood experiences, self-rated childhood health, childhood smoking, and menarche, adulthood factors: education, previous labour market participation, nulliparity, children under 18, illness, cohabiting partner, and accommodation type.
